# Human-to-human transmission of leptospirosis: A global systematic review

**DOI:** 10.1101/2025.09.02.25334635

**Authors:** Kaya Stollberg, Martin Richter, Ulrika Windahl, Olena Pyskun, Johanna F. Lindahl, Martin Wainaina

## Abstract

**Background:** Leptospirosis is an important zoonotic disease that is widely distributed globally. Transmission typically occurs when a person is exposed to infected animals or contaminated environments through broken skin or mucous membranes, either direct or indirectly through occupational or recreational activities. Cases of human-to-human transmission of leptospirosis are reported non-systematically, and their public health implications are currently unknown. We therefore aimed to review cases of transmission between persons documented in medical literature.

**Methodology:** Systematic literature searches were performed on Embase, PubMed, Scopus, and Web of Science databases to identify relevant publications without any time or language restrictions. Possible modes of human-to-human transmission, namely, aerosols, blood transfusion, breastfeeding, coitus, direct mucosal contact, parturition, and tissue transplantation were considered during literature searches. Clinical data describing the index and secondary cases were extracted from the included publications. The strength of the evidence of transmission between persons was assessed using the following criteria: (i) laboratory-confirmed infections in both index and secondary cases (or laboratory-confirmed infections in foetuses/neonates only), (ii) a clear epidemiological link, (iii) temporal alignment of both infections, and (iv) biological plausibility of the transmission route.

**Results:** The search yielded 8257 results, of which 27 publications covering 34 suspected reports from years 1932-2022 were finally included. Twelve of these 34 cases met all 4 quality criteria (strong evidence), five fulfilled 3 criteria (moderate evidence), and seventeen satisfied 1-2 criteria (weak evidence). Strong (n=12), moderate (n=3) and weak (n=15) evidence was found for vertical transmission, occurring either transplacentally (n=28) or via lactation (n=2). Moderate and weak evidence for transmission via coitus (n=3), and weak evidence via blood transfusion (n=1) was found. No reports of human-to-human transmission through organ transplantation or aerosols was found.

**Conclusion:** Human-to-human leptospirosis transmission may play a role in leptospiral epidemiology, especially through vertical spread. Public health systems, especially in endemic and outbreak-prone areas, should remain vigilant of these less-reported modes of transmission to safeguard maternal and neonatal health, transfusion safety and sexual well-being. Robust One Health surveillance is required to improve detection and awareness, reduce underreporting and improve disease control for better public health outcomes.

**Author summary:** Leptospirosis is an important neglected tropical disease that is distributed globally. Transmission to humans is commonly through infected animals (zoonotic) or via contact with contaminated soil and water. Extreme weather events such as flooding and heavy rainfall caused by climate change can drive increased leptospirosis outbreaks. Human-to-human transmission is less reported and thought to be epidemiologically unimportant. However, with the increased number of cases globally, these rare transmission modes may become more noticeable in certain contexts. We therefore conducted this systematic literature review to investigate clinical evidence of transmission of leptospirosis between persons from global medical literature. We found 27 reports spanning 90 years that covered 34 suspected cases from 17 countries in all continents except Antarctica. Reports revealed strong evidence for vertical transmission of leptospirosis (mother to child), and a possibility of transmission via sexual intercourse and blood transfusion. Boosting surveillance and diagnostics in endemic areas is recommended to reduce disease burden and improve health outcomes.

## Introduction

Leptospirosis is a globally important zoonotic disease with over one million reported human cases and 58,900 deaths annually [1]. It is caused by bacteria of the genus *Leptospira* (*L*.) [2], which commonly infect animal hosts that can shed the pathogen in their urine. Small mammals, especially rats and other rodents, are important reservoirs for the pathogen. Infection in humans occurs when bacteria gain entry through interrupted skin or mucous membranes. This can occur through direct or indirect contact with infected animals or contaminated soil or water either occupationally (e.g., slaughterhouse workers, veterinarians, paddy field farmers), or recreationally (e.g., swimming, kayaking) [3, 4]. Humans are accidental hosts and infections can vary from asymptomatic to acute, with incubation periods between 3 and 30 days. Fulminant forms that involve multiple organs, e.g., leptospirosis-associated pulmonary haemorrhage syndrome, meningitis/meningoencephalitis and Weil syndrome associated with jaundice, renal failure and myocarditis, can occur and often have high case fatality ratios [3, 5]. Leptospirosis is often underreported due to inadequate diagnostic capacity in endemic areas, and has a clinical presentation that often lacks distinguishing features from other endemic acute febrile illnesses [6, 7]. Diagnosis can be done directly using culture and PCR tests, or indirectly by detecting antibodies using serological tests such as the microscopic agglutination test (MAT) or ELISA [8].

There has been an increase in leptospirosis cases in various regions globally with time [9, 10]. Extreme weather events such as flooding and heavy rainfall are thought to be important disease drivers, alongside population growth and urbanisation [11]. Human-to-human transmission is a less reported mode of leptospiral transmission that has historically been given little attention in leptospiral epidemiology. However, with the dynamic nature of these disease drivers and risk factors for human infection, it remains unclear whether these rare modes of transmission could play more important roles in disease transmission in the future. We therefore examined the literature to determine documented cases of transmission between persons and discuss the implications of the evidence to strengthen preparedness in the face of a changing disease landscape.

## Methods

The literature search was conducted on 24^th^ September 2023 in leading databases which comprised Embase, PubMed, Scopus, and Web of Science. A review protocol was developed according to the Preferred Reporting Items for Systematic Reviews and Meta-Analyses (PRISMA) guidelines [12]. The review was registered on OSF registries [13]. The following search terms connected with Boolean operators were applied; (leptospira OR leptospirosis) AND (transfusion OR transplant OR vertical OR pregnan* OR human-to-human OR person-to-person OR sexual OR lactation OR aerosol OR mucosa) (S1 Data). No limitations on language or publication time were placed in the searches.

The citations were imported into Endnote 21 (Thomson Reuters, Philadelphia, PA, USA) and duplicates were removed. All citations were screened using the titles and abstracts for relevance, and the DeepL translation tool [14] was used for non-English text. For initial screening, titles and abstracts were divided among all six authors, ensuring each record was independently reviewed by a second author. Discordant results were then resolved by MW. Two authors (KS and MW) conducted the data extraction from included publications.

Peer-reviewed journal articles that reported possible transmission between humans were included. In cases of suspected vertical transmission but with no laboratory diagnosis, reports were excluded if it was clear that foetuses died from complications of maternal leptospirosis, or had positive outcomes with no indication of illness. Studies that reported leptospirosis in either index or secondary cases with the biological plausibility of transmission between persons were retained. Serological proof without accompanying clinical presentations consistent with leptospirosis were rejected as they were too speculative of potential transmission between people. The articles were screened independently by KS and MW and disagreements were discussed and resolved with co-authors.

Quality assessment was done according to a tailor-made tool designed to assess the strength of evidence for human-to-human transmission (Table 1) and summarised using the *robvis* tool [15].

**Table 1.** A custom-made quality assessment tool used to gauge the strength of evidence for human-to-human transmission of leptospirosis.

| Criteria | Criteria matched | Level of evidence | Interpretation |
| --- | --- | --- | --- |
| Domain 1 (D1): Laboratory-confirmed infection in both index and secondary cases* | 4 | Strong | High quality evidence of transmission between persons |
| Domain 2 (D2): Clear epidemiological link | 3 | Moderate | Probable transmission but with incomplete evidence |
| Domain 3 (D3): Temporal alignment between the two infections | 0-2 | Weak | Anecdotal indication of transmission |
| Domain 4 (D4): Transmission route is biologically plausible |  |  |  |

\* In cases of vertical transmission, laboratory-confirmed infections in foetuses or neonates ( $\leq 1$ month) were enough to fulfil this criterion as environmental or zoonotic exposure was considered unlikely.

## Results

### Database search results

A total of 8257 references were identified in the primary search, with Scopus giving 6085 hits, and Embase (n=996), PubMed (n=779), and Web of Science (n=397) following in decreasing order. The removal of duplicates from the combined search results gave 6326 remaining references which were screened for relevance by title and abstract. Ultimately, 27 studies comprising 34 reports were considered relevant for this review (Fig 1).

**Fig 1.**
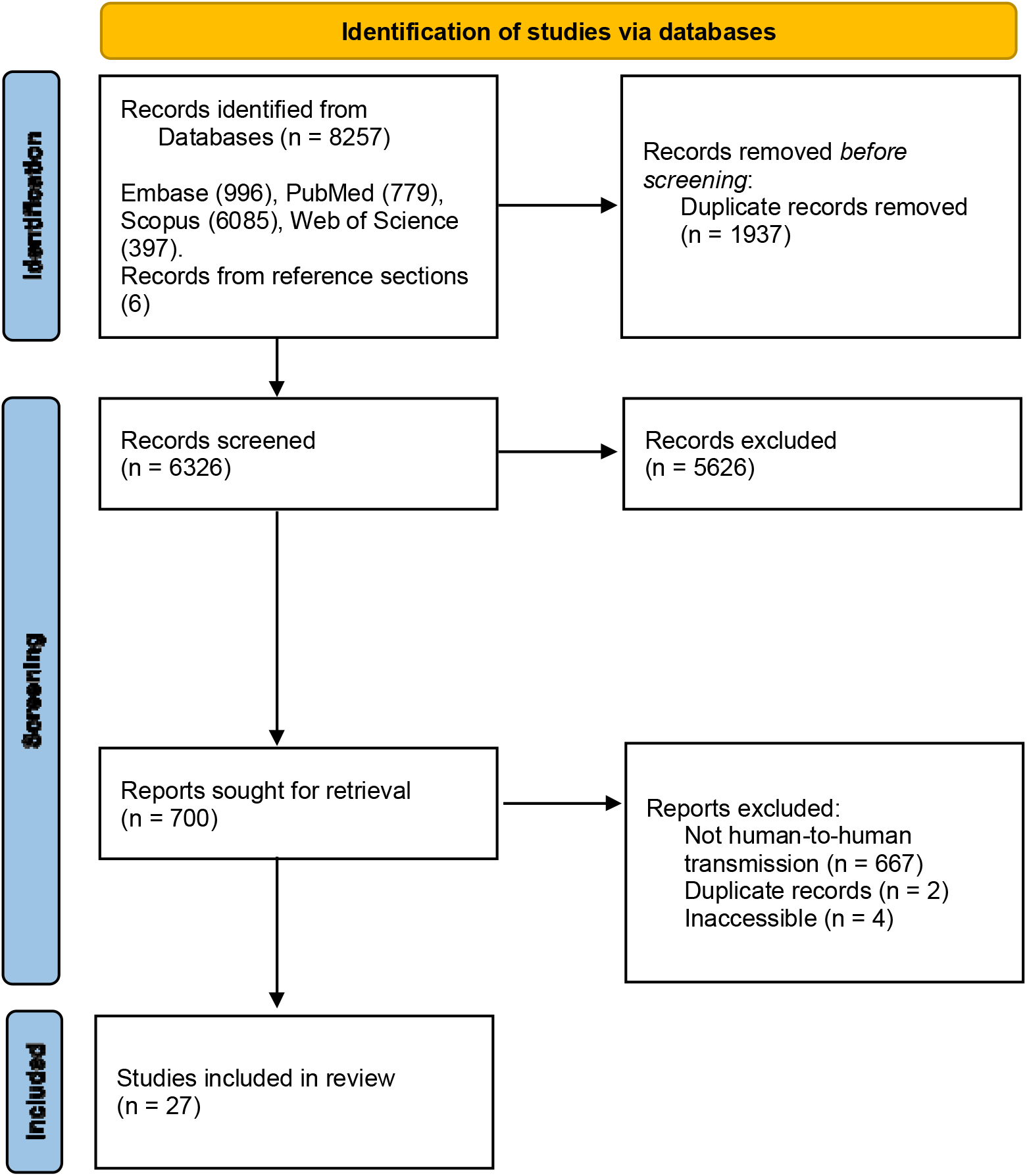
The study selection process utilised to identify relevant literature on human-to-human transmission of leptospirosis from leading electronic databases

### Study characteristics

The studies were published between 1932 and 2022, with most being in English (17) and the rest in French (3), German (3), Chinese (1), Dutch (1), Hungarian (1) and Spanish (1). Most publications were from Europe (10), followed by Asia (8), South America (4), Africa (2), North America (2) and Oceania (1) in decreasing order. Lastly, most publications were case reports/series (22), while cross-sectional (4) and case-control (1) studies were fewer (S2 Data).

### Quality assessment of evidence

We assessed the strength of the evidence for the transmission of leptospires between persons and revealed that overall, 12 reports (35.3%) had strong evidence of transmission, and 5 (14.7%) had moderate and 17 (50.0%) showed weak evidence (Fig 2).

**Fig 2.**
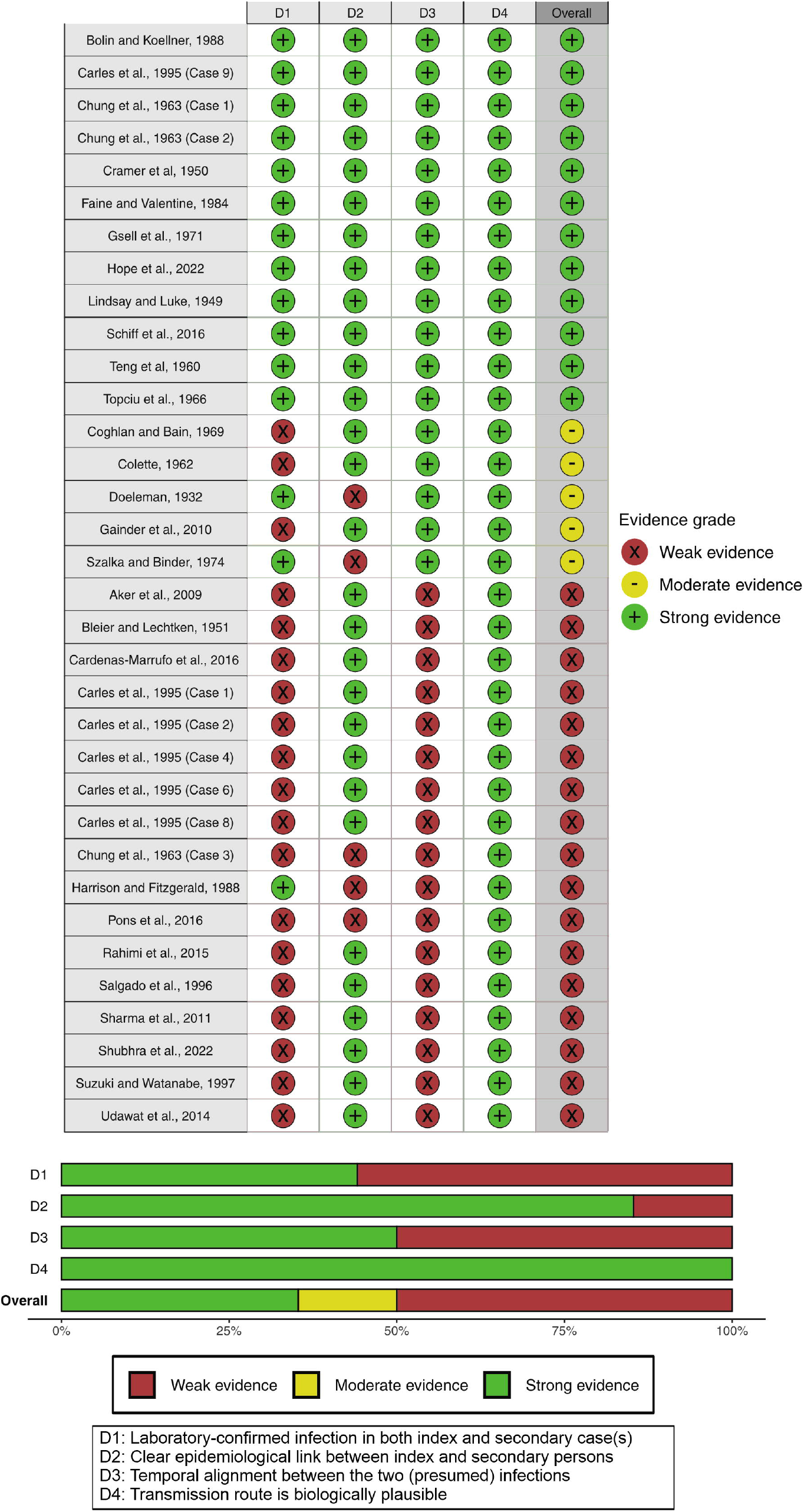
Assessment scores of the strength of evidence for transmission between persons based on four criteria from 34 reports. A summary bar chart is also presented with the results categorised into weak, moderate and strong evidence of transmission.

### Vertical transmission

Transmission from mother to child was indicated in 30 studies, among which 12 studies showed strong, 3 had moderate and 15 demonstrated weak evidence of transmission (Table 2, Fig 2). Most cases in the infant were acquired *in utero*, resulting in stillbirths/intrauterine death, and some cases were carried to term and resulted in neonatal death, or healthy/mildly affected neonates (Fig 3). Two cases were reportedly transmitted via breastfeeding, and were backed by strong evidence where leptospires were detected from both maternal and neonatal samples, and weak evidence where leptospires were detected in breastmilk but without neonatal testing. Sepsis was a commonly reported clinical manifestation in the affected neonates. Two studies lacked testing from maternal samples (Table 2), but *Leptospira* IgM antibodies and DNA were found in the neonates, indicating strong evidence of vertical transmission.

**Table 2.**
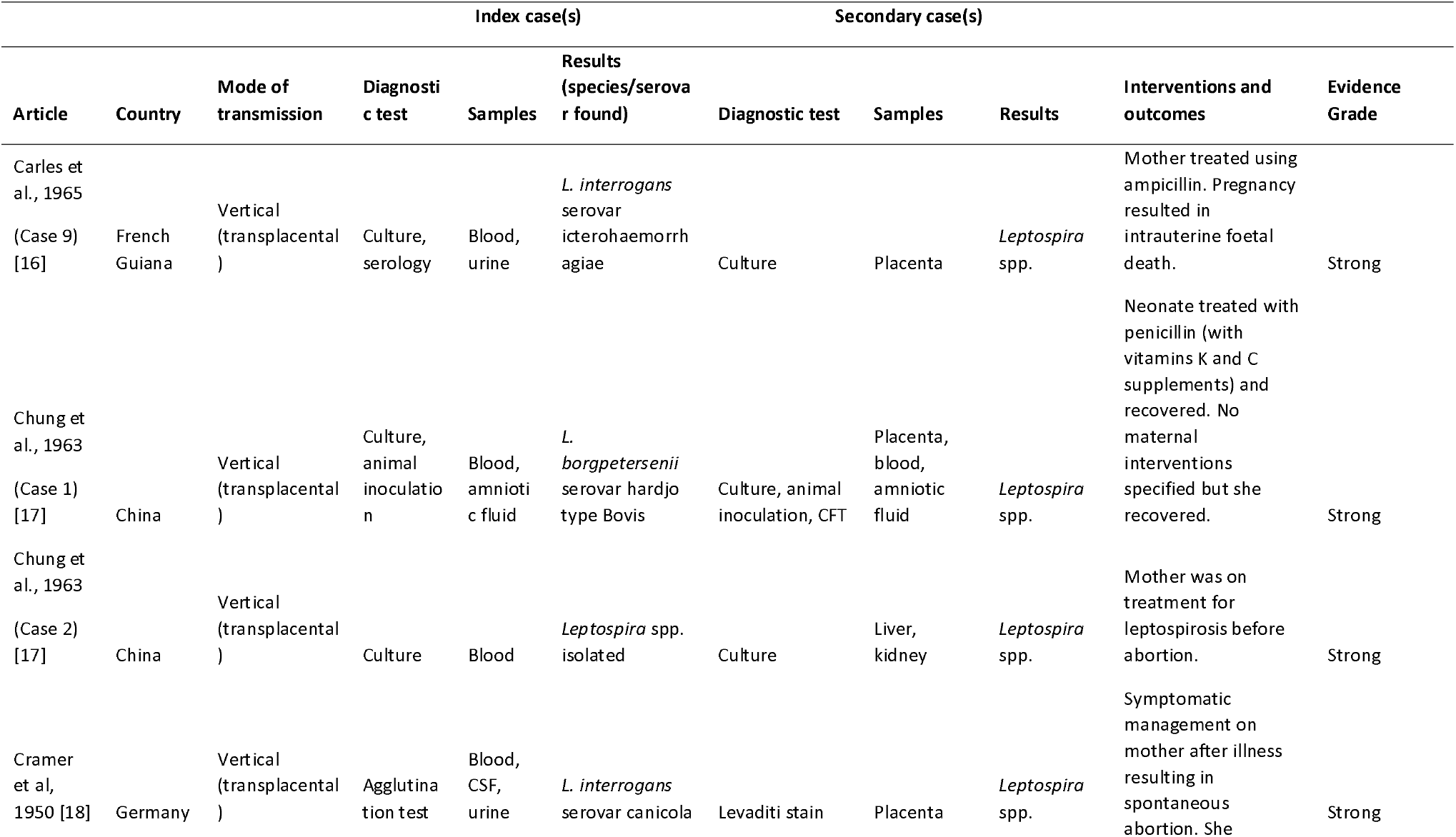
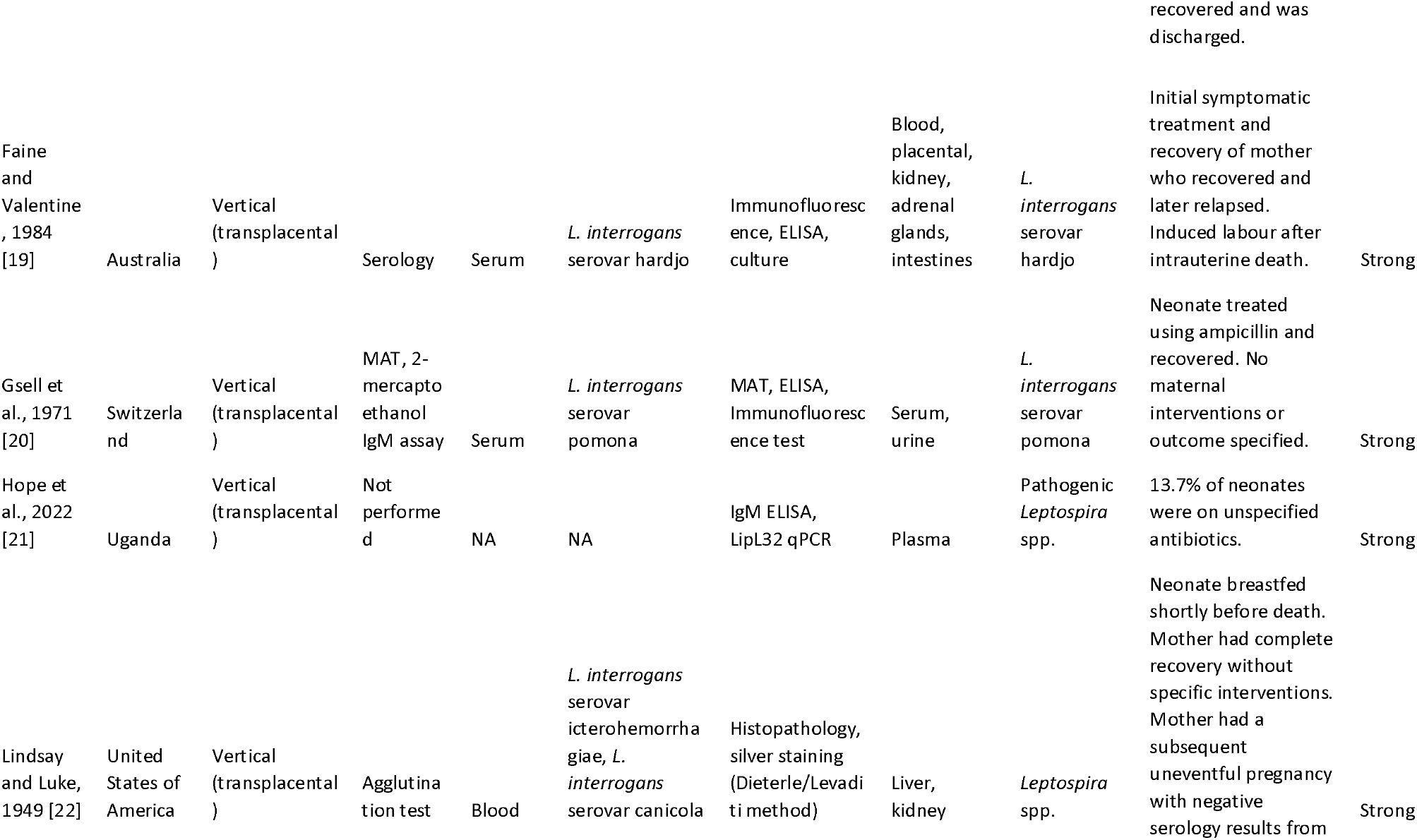
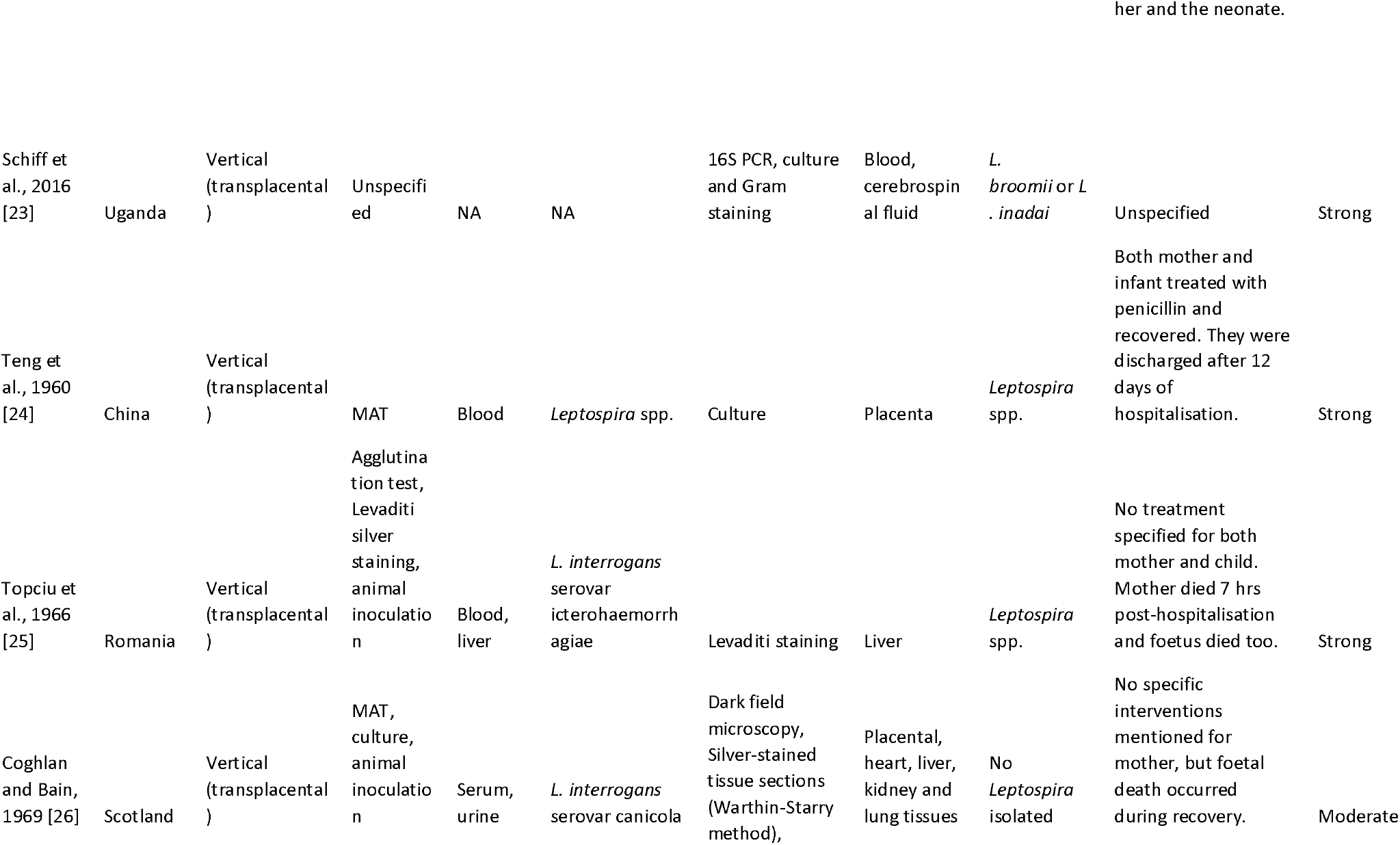
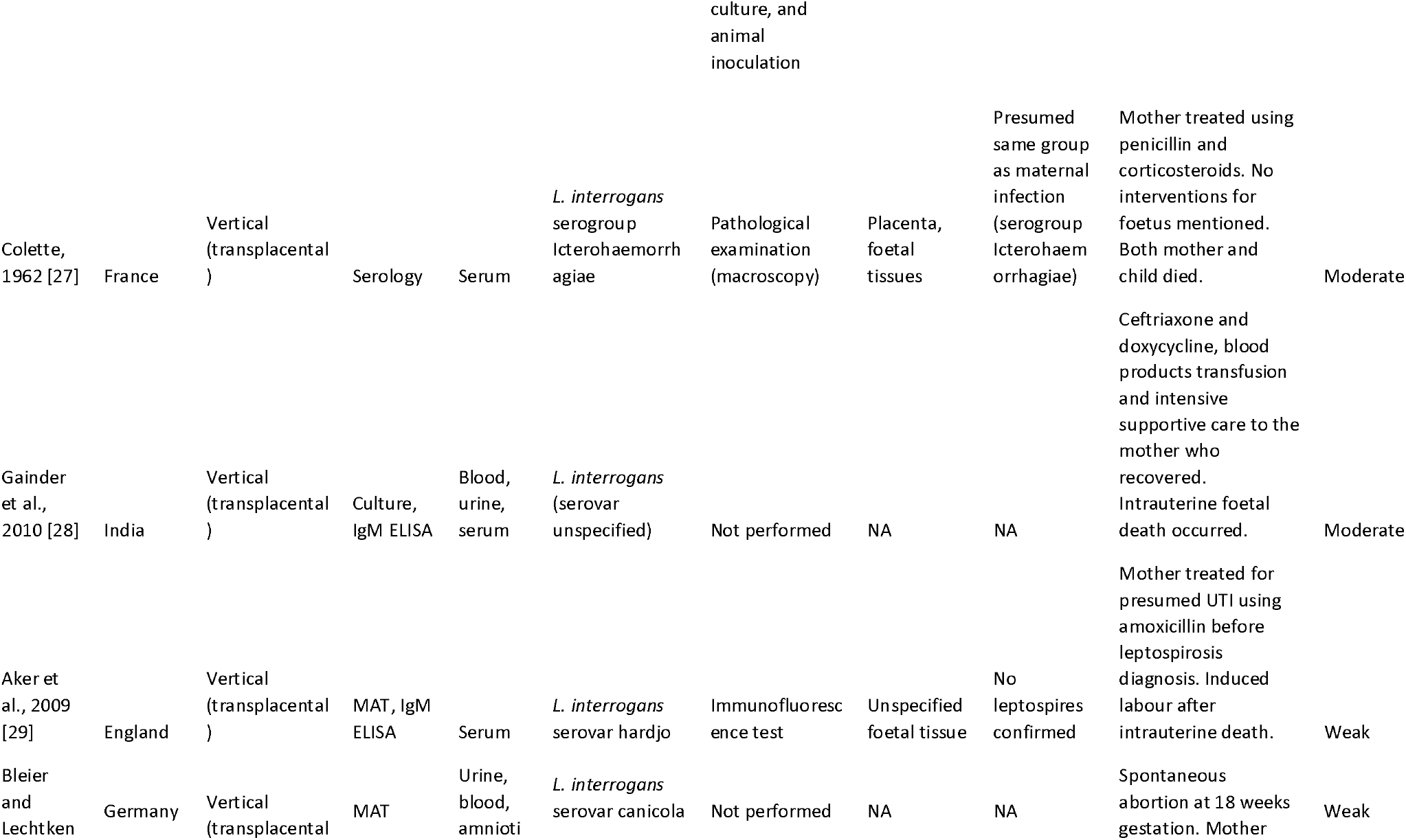
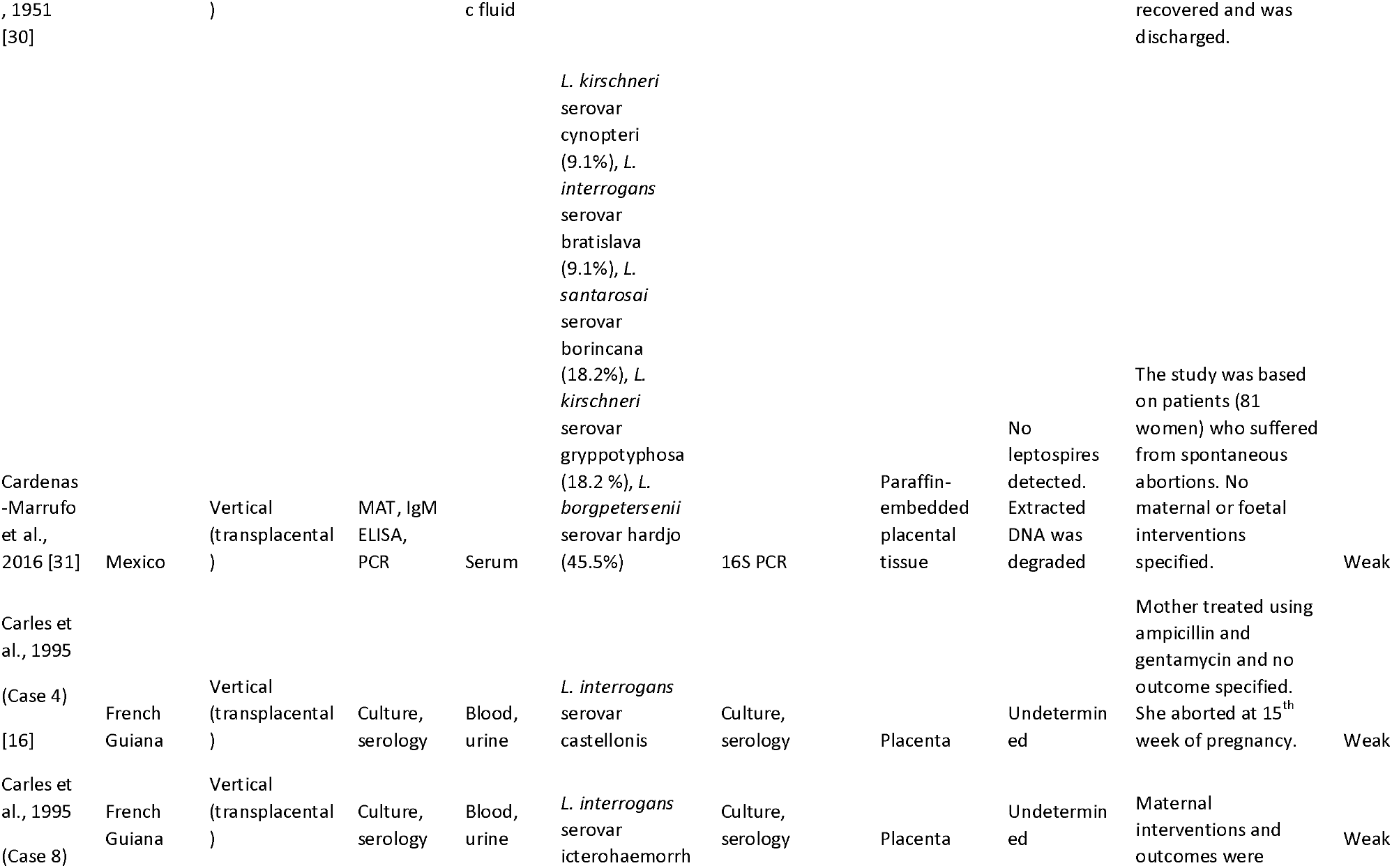
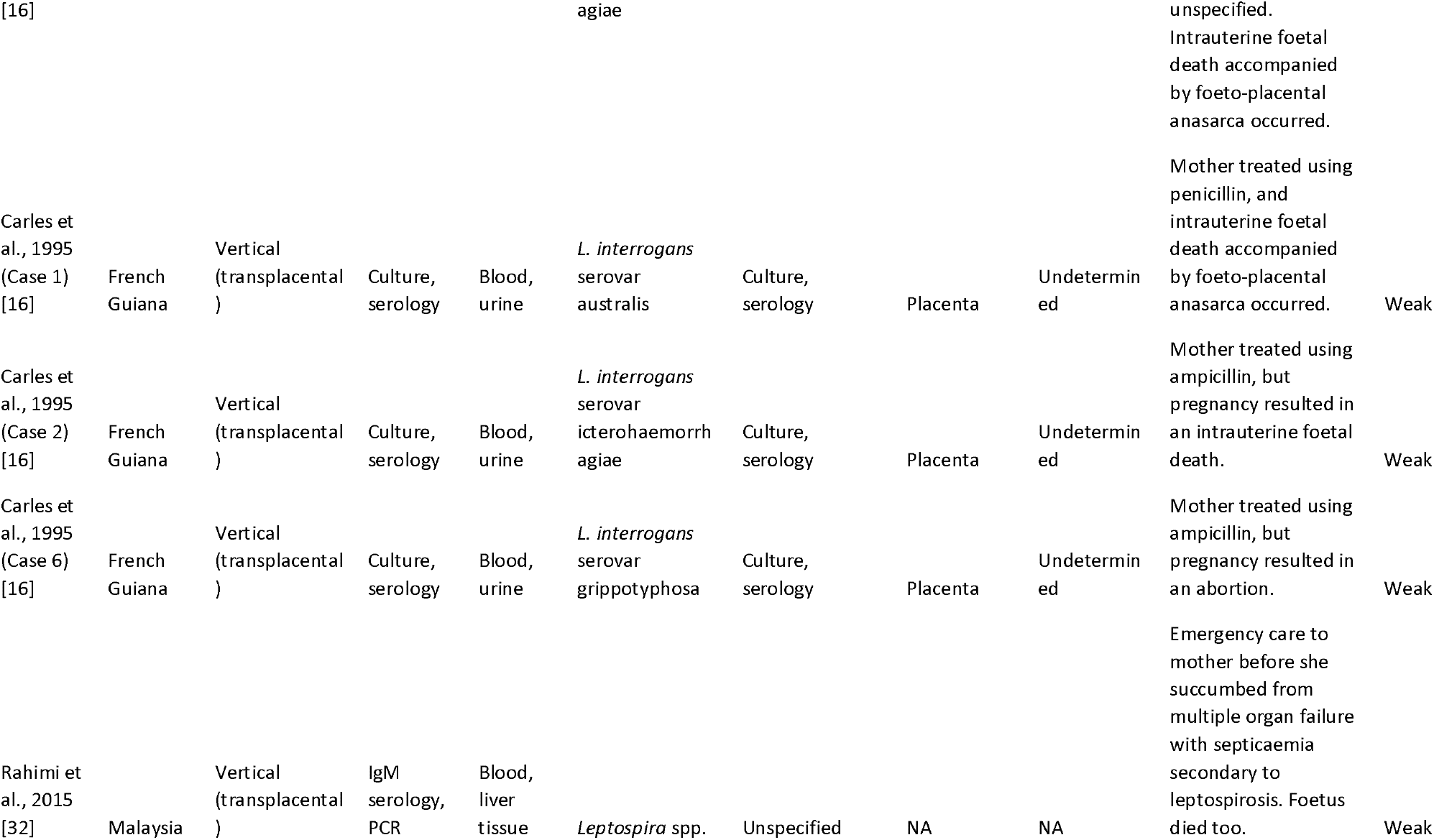
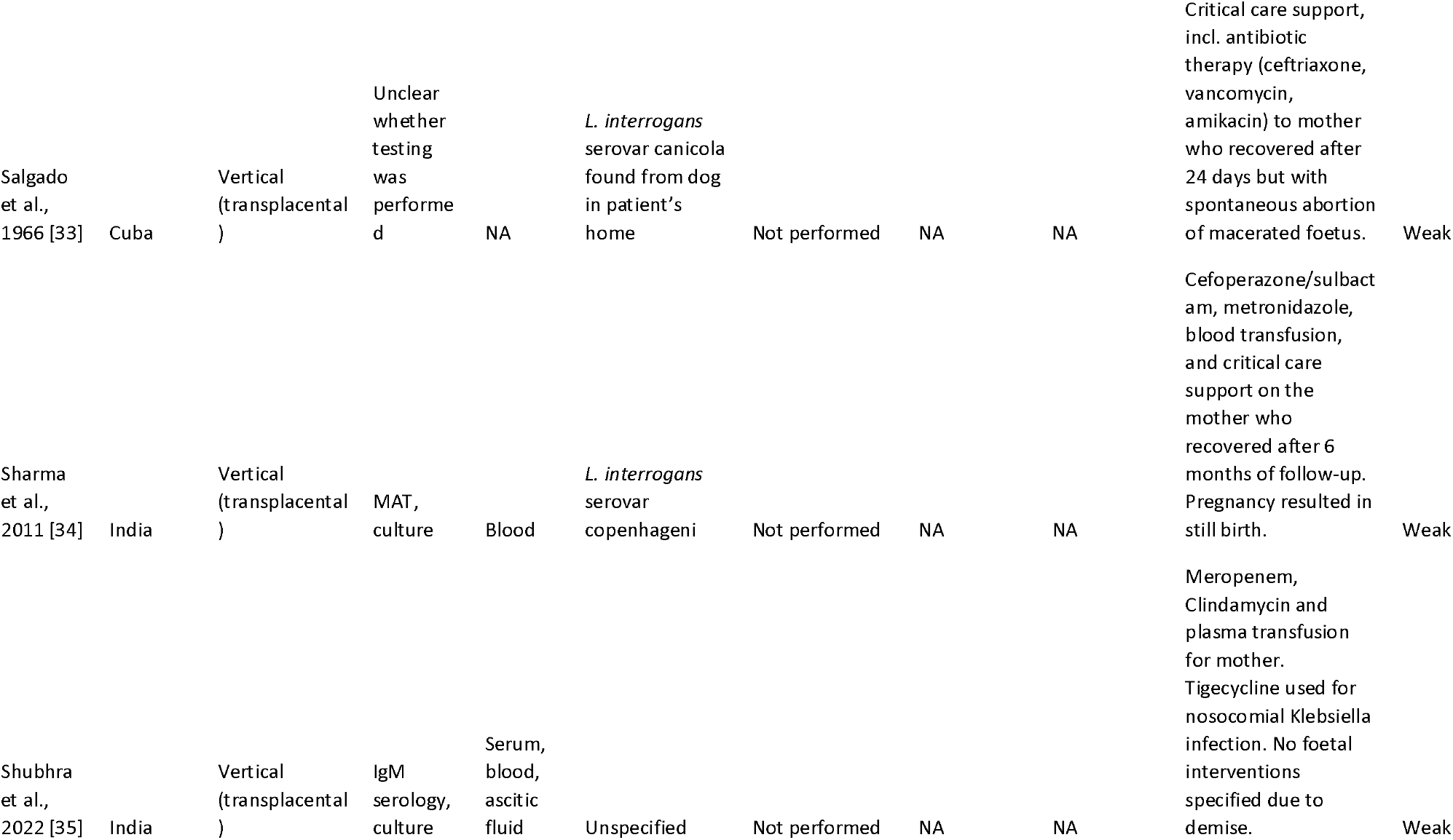
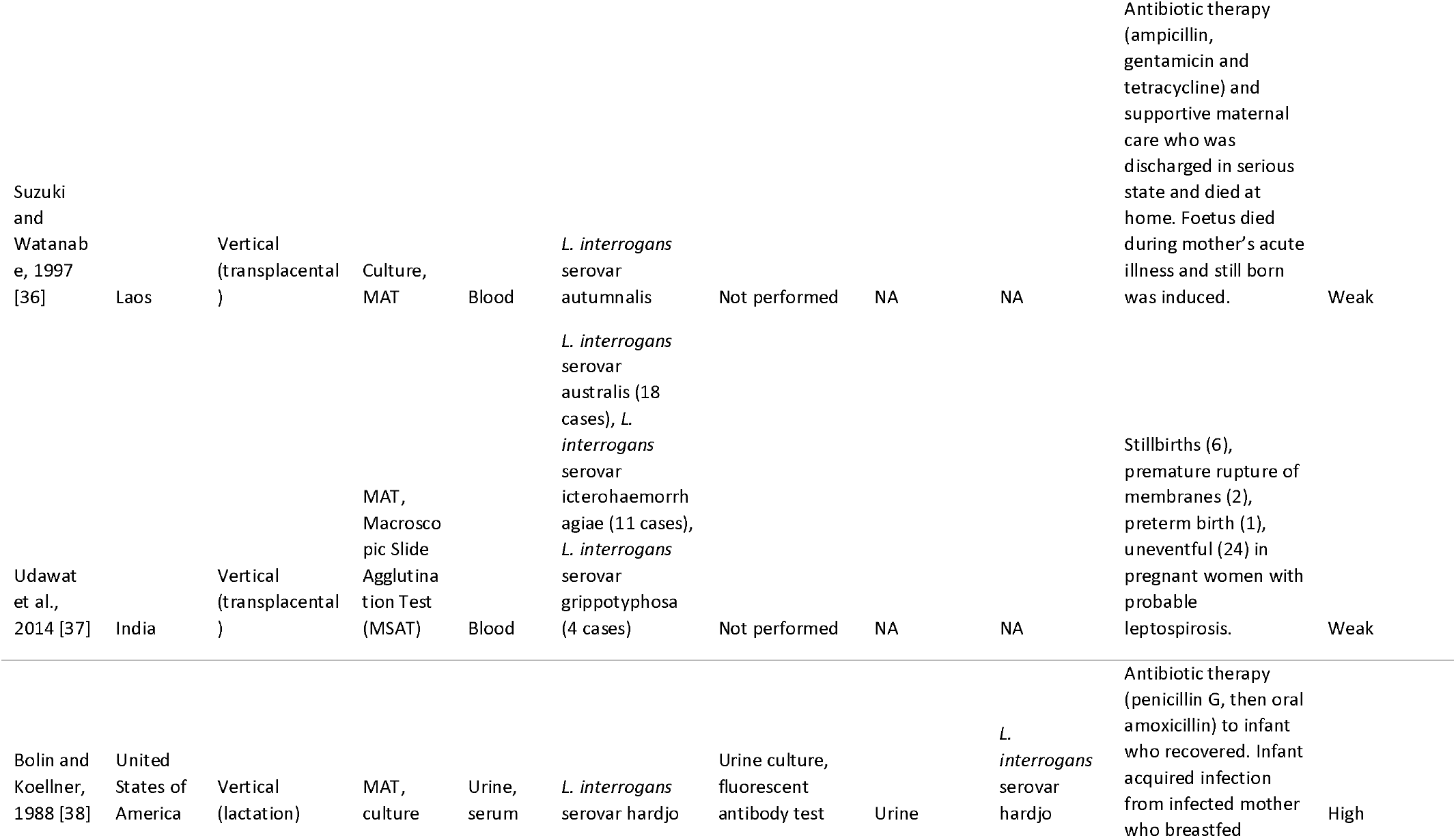
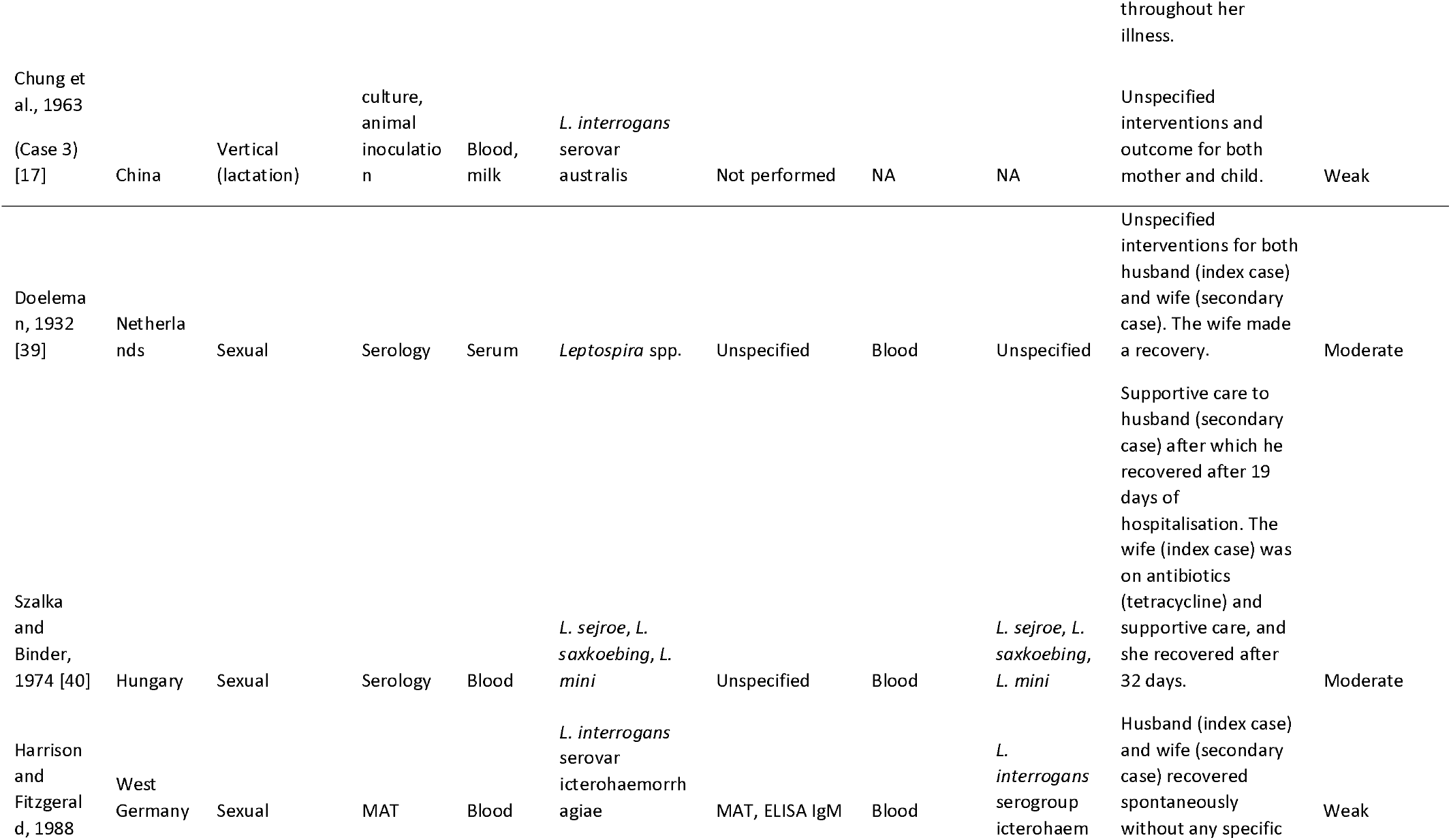
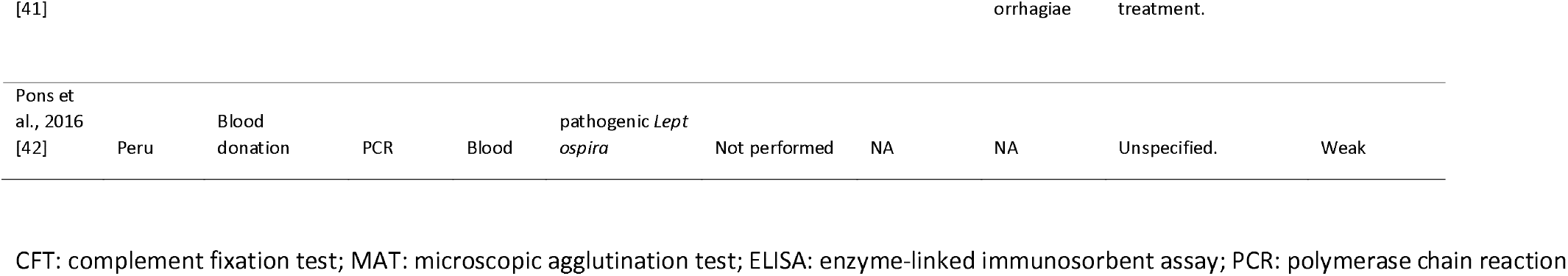
A summary of study characteristics of publications investigating human-to-human transmission of leptospirosis globally. Studies are presented according to the mode of transmission and evidence grade determined.

**Fig 3.**
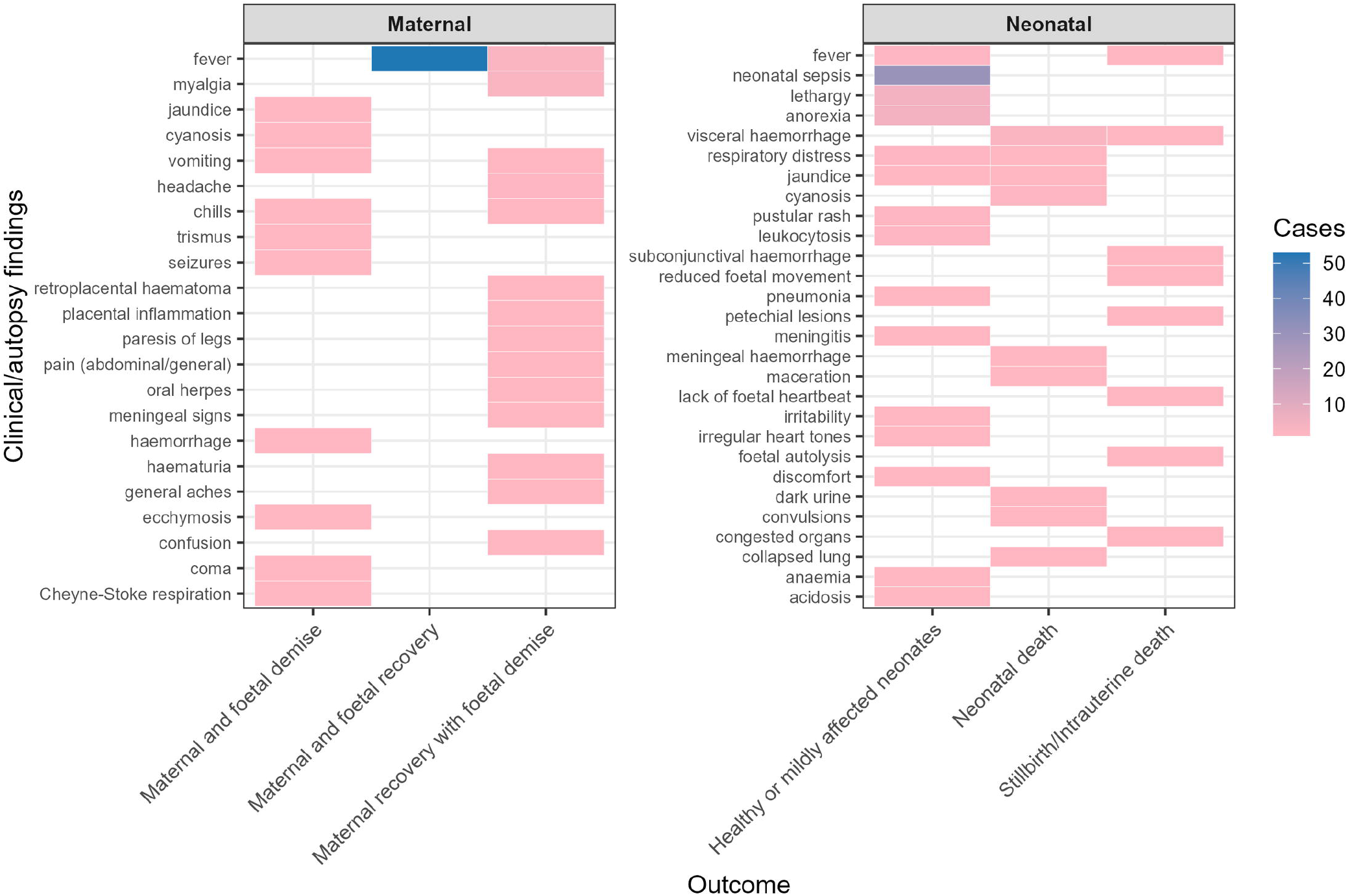
Heatmaps of clinical findings in infants and mothers with disease outcomes of cases of vertical leptospiral transmission with strong evidence.

Additionally, maternal clinical findings were diverse and resulted in maternal recovery with foetal death or recovery (Fig 3). Fever, jaundice, myalgia and abdominal pain were common clinical presentations in infected mothers.

Laboratory confirmation was done using the MAT, ELISA, PCR, culture, immunohistochemistry tests, on maternal, infant, or paired maternal-neonate samples (Table 2). Where no laboratory testing was done on either index or secondary cases, clinical presentations or autopsy findings were relied upon to suspect transmission.

Putative risk factors associated with maternal exposure were occupational exposure to infected livestock through veterinary practice and farming, contact with infected pets, travel to endemic regions, farming in paddy fields, living near water/lowlands, exposure to rodent reservoirs, and swimming (S2 data).

### Sexual transmission

Three dated case reports were found reporting sexual transmission of leptospirosis between heterosexual partners, and the evidence was of moderate to weak strength (Fig 2). Recreational exposure was a common risk factor in the index cases, and diagnosis was performed serologically using MAT and ELISA. Therefore, while sexual transmission was posited in the three reports, it was not proven by microbiological isolation of sexual fluids, bacterial typing beyond serovar identification, and robust contact tracing to ensure ruling out alternative sources of infection.

### Blood donation

A cross-sectional study was found demonstrating pathogenic/intermediate leptospires via 16rRNA (*rrs* gene) PCR from 8/42 (19%) healthy blood donors in Cajamarca, Peru (Table 2). The area was described as rural with farming and exposure to rodent carriers as possible risk factors, and donors who were at risk of environmental and occupational exposure were included in the study, and thus not reflective of all blood donors. No transmission was established in blood recipients, but the demonstration of DNA in asymptomatic donors suggest subclinical carriage of the bacteria. The evidence for transmission was graded as weak (Table 2).

### Other transmission modes

No evidence was found for transmission between persons via aerosols and organ transplant.

## Discussion

We reviewed global literature spanning more than 90 years to investigate evidence of leptospirosis transmission between humans, a less reported route. Strong evidence was found for transmission from mother to child either via the placenta or breastfeeding. Moderate to weak evidence for sexual transmission was observed and weak evidence was found for transmission through blood transfusion. We discuss the implications of these biologically plausible routes to current public health policy and call for future studies with stronger evidence to better understand the risk of transmission and the impacts on disease prevention and control.

Entry of *Leptospira* during infection typically occurs through the interrupted skin and mucous membranes. Direct contact with the biological fluids of an infected animal such as urine, or indirect exposure to contaminated soil or water are responsible for most cases. Transmission between persons is biologically plausible via blood transfusion, tissue/organ transplant, vertically (during pregnancy or lactation), through sexual contact, through aerosols/droplets. However, sustained human-to-human transmission of zoonoses depends on the transmission route (e.g., airborne, contact, faecal-oral, and vector-borne transmission), host, pathogen, and environment [43, 44]. Additionally, characteristics of the index and secondary persons, (e.g., immune function, infectious dose), and transmission amplifiers that act between the index and secondary cases (e.g., climate, human behaviour, cultural variation) are important [44].

### Vertical transmission

Vertical transmission generally refers to the transfer of an aetiological agent from either parent to offspring, occurring mostly via the mother across the placenta (*in utero*), during childbirth (intrapartum), or after birth (postpartum) by breast-feeding [45]. In our case, we observed mother-to-child transmission of leptospirosis occurring *in utero* (transplacentally) in most instances, with one case occurring via lactation with strong evidence. Vertical transmission has been demonstrated in some domestic animals, suggesting its likely importance in human hosts [46–50]. Most publications were case reports or case series, highlighting the case-based nature of this transmission mode of leptospirosis, and need for stronger evidence through superior study designs with more statistical power [51]. Still births and intrauterine deaths, congenital abnormalities (e.g., hypotrophy), low birth weight, and small for gestational age were observed in neonates of leptospirosis-positive mothers. Fever, anaemia, jaundice and myalgia, as well as severe outcomes such as maternal mortality and pregnancy loss were common features of maternal leptospirosis, and were consistent with a recent comprehensive review of leptospirosis during pregnancy [52] where cases were also acquired from environmental and zoonotic sources. Proving leptospirosis transmission during human gestation is not always conclusive, especially when there is no foetal/neonatal testing, a lack of obvious adverse foetal outcomes such as illness or death [53–61], or foetal death occurred from maternal complications from leptospirosis or otherwise [62–64]. Therefore, it is likely our review introduced a selection bias by excluding unclear cases of vertical transmission which may underrepresent the real-world limitations of leptospirosis diagnostics. Additionally, histological proof of leptospires after foetal death was not always successful due to tissue maceration and likely death of leptospires, which can render test results inconclusive [26, 27]. However, proper treatment and patient management is recommended as vertical transmission is likely unreported, especially in endemic areas, and a likely source of reporting bias in this review.

Lactation has been implicated as a possible transmission route in this review [38], a phenomenon observed in animal models too [65]. The Chinese Medical Association has recommended the pasteurisation of breastmilk from infected mothers during treatment as it remains more beneficial to the infant than formula milk. Clinical suspicion of leptospirosis in endemic areas is critical, and boosting surveillance and diagnostics in perinatal care is important in reducing disease burden which is likely underreported.

### Sexual transmission

Diseases are sexually transmissible when the aetiological agent can be passed among persons via bodily fluids such as blood, semen and vaginal fluids during oral, anal, or genital sex with an infected partner [66]. Additionally, categorising diseases as Sexually Transmissible Infections (STIs) requires sexual contact as the primary mode of transmission [66]. Epidemiological comparisons of disease rates by sex, and molecular cluster analyses among sexually-linked samples can help recognise emerging STIs [67]. The three studies positing sexual transmission of leptospirosis [39–41] were published between 1932-1988, when identifying leptospires largely relied on serology and culture methods, and no PCR assays were available yet [68]. Consequently, no leptospires were demonstrated in the patients’ bodily fluids associated with sexual transmission, and only circumstantial evidence was provided, e.g., acute disease after sexual contact with partner with known exposure to partner with no discernible risk factor. This moderate evidence makes us conclude that sexual transmission of leptospirosis is currently not well proven. Studies are required to demonstrate leptospires and their survival in semen, vaginal fluids, genital and anal swabs, etc. from infected persons, and molecular analyses showing the phylogenetic relationship between linked cases are required to raise the understanding on this transmission mode. Providing detailed exposure history through contact tracing could help track cases, but using this approach is limited in STIs due to its associated intrusive nature and stigmatisation [69]. Ruling out other sources of exposure by animal and environmental sampling is critical in strengthening evidence of sexual transmission of leptospirosis. However, venereal leptospiral transmission has been observed in various domestic animals and murine models [48, 70–73], and is therefore biologically plausible in humans.

### Transfusion-mediated transmission

Various emerging and re-emerging pathogens that are not part of standard screening pose a risk to transfusion medicine [74]. The included study demonstrated leptospiral DNA in donated blood from high-risk donors in an endemic area in Peru, offering credible evidence of potential transmission [42]. Several studies demonstrating past infections in blood donors using serology also exist, showing the potential risk of transmission via blood transfusion [75–78]. Bacteriaemia is often associated with the initial leptospiraemic phase during the first 8 days of fever [3], but PCR positivity in asymptomatic blood donors suggest subclinical bacterial carriage, the risk of transmission of which is not clear. Eligibility criteria for blood donation that filter out febrile syndrome would do well in curbing transmission via transfusion of blood and blood products. However, blood donor eligibility varies substantially across nations, and criteria that are evidence-based are recommended [79]. Pathogen reduction technologies for donated blood such as irradiation could also play a role in reducing transmission of emerging pathogens [74]. Artificial oxygen carriers (artificial blood) are novel approaches that may help prevent transmission of emerging transfusion-mediated infections, especially in patients requiring repeat transfusions such as sickle cell anaemia and thalassemia. However, these therapies are not yet approved for use in several countries due to high toxicities [80, 81].

### Organ transplant-mediated transmission

No reports of transmission via organ transplants were found. However, solid organ transplant guidelines in the endemic south Asian region have recommended leptospirosis screening in organ donors and recipients with significant epidemiological risk or exposure due to the possibility of transplant-mediated leptospirosis [82]. Additionally, donation should be deferred to 3 months after recovery, and treatment in recipients be done with as in immunosuppressed patients [82]. Case reports of liver and kidney transplant recipients who acquired leptospirosis due to immunosuppressed states have been reported, and the infections were attributed to contaminated environments and not donor-derived [83, 84].

### Aerosol transmission

No reports of airborne transmission were found. Pulmonary involvement in leptospirosis is well documented, and characterised by chest pain, cough, dyspnoea and haemoptysis. Acute respiratory distress syndrome (ARDS) and severe pulmonary haemorrhagic syndrome (SPHS) are severe forms of leptospirosis that can lead to respiratory insufficiency and death [85, 86]. Leptospires have also been detected in respiratory samples of patients with sudden worsening of chronic obstructive pulmonary disease (COPD) by amplicon sequencing of endotracheal aspirates [87]. Hamster models have shown that leptospires have predilection for lungs as evidenced by pulmonary lesions, haemorrhage, and elevated inflammation markers after experimental inoculation [88]. Consequently, the possibility of transmission via droplets from ill patients to surrounding persons cannot be ruled out.

## Conclusion

We provide a comprehensive synthesis of reported cases of human-to-human transmission of leptospirosis. Despite exposure to contaminated environments and infected animals remaining the most important ways of acquiring leptospirosis in humans, credible evidence of vertical transmission, both transplacentally and via breastfeeding, was found. Evidence for sexual transmission was moderate to weak. Possible risk of transmission through blood-transfusion was found (weak evidence), and no reports of transmission via solid organ transplantation and aerosols between persons was found, despite being biologically plausible. These findings demonstrate the need to consider leptospirosis in maternal and neonatal healthcare, especially in endemic and outbreak-prone areas. As risk factors for exposure to the index cases of infections found in this review were zoonotic or environmental, robust surveillance systems that utilise integrated One Health approaches are required. Applying molecular diagnostics to determine the traceability of cases will help elucidate cases of transmission of leptospirosis between persons and better understand the risks posed to human health, especially in transplant and transfusion medicine, sexual wellbeing, and maternal and neonatal health.

## Data Availability

All data produced in the present work are contained in the manuscript

## Declarations

### Ethical approval

Not applicable

### Data availability

All data related to this study are included in this published article and its supplementary information files.

### Author contributions

**Kaya Stollberg** (Methodology, Formal analysis, Validation, Investigation, Data curation, Writing - Original draft, Writing - Reviewing); **Martin Richter** (Investigation, Writing - Reviewing); **Olena Pyskun** (Investigation, Writing - Reviewing); **Ulrika Windahl** (Investigation, Validation, Writing - Reviewing); **Johanna F. Lindahl** (Methodology, Investigation, Validation, Writing - Reviewing); **Martin Wainaina** (Conceptualisation, Data curation, Formal analysis, Investigation, Methodology, Project administration, Supervision, Validation, Visualisation, Writing - original draft, Writing - review & editing)

## Acknowledgements

We thank Dr. Jiaxin Ling (Uppsala University) for translating the traditional Chinese text. We also thank the authors of the included reports.

## Supporting information legends

S1 Data: Database search terms used to find literature on leading electronic databases. The PRISMA checklist is also given presented.

S2 Data: A summary of the data extracted from the included studies.

