## Supplementary material for "Human-to-human transmission of leptospirosis: A global systematic review": S1 Data

Literature searches were conducted on 24.09.2023.

Table 1: A summary of the search terms used for literature identification

| **Search terms (All fields where setting is available)** | **Database** | **Results** |
| --- | --- | --- |
| (leptospira OR leptospirosis) AND (transfusion OR transplant OR vertical OR pregnan* OR human-to-human OR person-to-person OR sexual OR lactation OR aerosol OR mucosa) | PubMed | 779 |
| (leptospira OR leptospirosis) AND (transfusion OR transplant OR vertical OR pregnan* OR human-to-human OR person-to-person OR sexual OR lactation OR aerosol OR mucosa) | Scopus | 6085 |
| ('leptospira'/exp OR leptospira OR 'leptospirosis'/exp OR leptospirosis) AND (transfusion OR transplant OR vertical OR pregnan* OR 'human to human' OR 'person to person' OR sexual OR lactation OR aerosol OR mucosa) | Embase | 996 |
| (leptospira OR leptospirosis) AND (transfusion OR transplant OR vertical OR pregnan* OR human-to-human OR person-to-person OR sexual OR lactation OR aerosol OR mucosa)(All Fields) | Web of Science | 397 |
| **Total** |  | **8257** |
